# Heterozygous variants in *KCNC2* cause a broad spectrum of epilepsy phenotypes associated with characteristic functional alterations

**DOI:** 10.1101/2021.05.21.21257099

**Authors:** Niklas Schwarz, Simone Seiffert, Manuela Pendziwiat, Annika Rademacher, Tobias Brünger, Ulrike B.S. Hedrich, Paul B Augustijn, Hartmut Baier, Allan Bayat, Francesca Bisulli, Russell J Buono, Ben Zeev Bruria, Michael G Doyle, Renzo Guerrini, Gali Heimer, Michele Iacomino, Hugh Kearney, Karl Martin Klein, Ioanna Kousiappa, Wolfram S. Kunz, Holger Lerche, Laura Licchetta, Ebba Lohmann, Raffaella Minardi, Marie McDonald, Sarah Montgomery, Lejla Mulahasanovic, Renske Oegema, Barel Ortal, Savvas S. Papacostas, Francesca Ragona, Tiziana Granata, Philipp S. Reif, Felix Rosenow, Annick Rothschild, Paolo Scudieri, Pasquale Striano, Paolo Tinuper, George A. Tanteles, Annalisa Vetro, Felix Zahnert, Federico Zara, Dennis Lal, Patrick May, Hiltrud Muhle, Ingo Helbig, Yvonne Weber

## Abstract

**Background:** *KCNC2* encodes a member of the shaw-related voltage-gated potassium channel family (K_V_3.2), which are important for sustained high-frequency firing and optimized energy efficiency of action potentials in the brain.

**Methods:** Individuals with *KCNC2* variants detected by exome sequencing were selected for clinical, further genetic and functional analysis. The cases were referred through clinical and research collaborations in our study. Four *de novo* variants were examined electrophysiologically in *Xenopus laevis* oocytes.

**Results:** We identified novel *KCNC2* variants in 27 patients with various forms of epilepsy. Functional analysis demonstrated gain-of-function in severe and loss-of-function in milder phenotypes as the underlying pathomechanisms with specific response to valproic acid.

**Conclusion:** These findings implicate *KCNC2* as a novel causative gene for epilepsy emphasizing the critical role of K_V_3.2 in the regulation of brain excitability with an interesting genotype-phenotype correlation and a potential concept for precision medicine.

## Introduction

Epilepsy is one of the most prevalent neurological diseases and affects approximately 50 million people worldwide^1^. Especially in children and young adults, epilepsy causes the most substantial burden compared to any other neurological condition^2^. The identification of epilepsy associated genes in the last decade has dramatically improved the understanding of epileptogenesis. More than ten epilepsy-associated genes code for potassium channels, partially leading to pathophysiology-based treatment, like the potassium channel blocker 4-aminopyridin in *KCNA2* caused developmental and epileptic encephalopathies (DEE)^3^. Based on the original description in *Drosophila melanogaster* potassium channels are devided in Shaker, Shaw, Shal and Shap subtypes^4^. The Shaw-related potassium channel family (K_V_3) play a pivotal role in the excitability of the central nervous system (CNS) by regulating the membrane resting potential, the firing of neurons, the action potential duration and neurotransmitter release^5,6^. So far, only *KCNC1* (K_V_3.1) and *KCNC3* (K_V_3.3) as members of this potassium channel gene family have been implicated in human neurological diseases. Disease-causing variants had previously been identified in patients with progressive myoclonus epilepsy and spinocerebellar ataxia^7,8^. *KCNC2* codes for the potassium channel K_V_3.2 (an additional member of the K_V_3 family) mainly expressed in the brain in the interneurons of cortex, hippocampus and basal ganglia^5^.

Here, in this study, we identified 25 different heterozygous variants in *KCNC2* in 27 unrelated individuals with developmental and epileptic encephalopathies (DEE) and other, more mild epilepsy syndromes such as genetic generalized epilepsy (GGE), early-onset absence epilepsy (EOAE), epilepsy with focal seizures (FE) and pure febrile seizures (FS) and provide a detailed phenotypical, genetic and functional analysis emphasizing the role of *KCNC2* as a novel disease gene in human epilepsies.

## Subjects and Methods

### Patients

We selected individuals with *KCNC2* variants, identified by exome sequencing, referred through clinical and research collaborations in our study, including individuals contributing through the Epi25 consortium^9^. Segregation analysis was performed when possible. No other relevant variants were detected in these cases based on the classification criteria by ACMG (American College of Medical Genetics)^10^. Clinical, neuroimaging and electrophysiological data were reviewed in detail. The current study was approved by each local ethics committee. The epilepsy syndromes were classified using the actual ILAE (International League against Epilepsy) criteria^11^. We hereby state that we received written informed consent to perform this study by each patient/relative included in the study.

## Methods

### Sanger sequencing analysis

We performed bidirectional Sanger sequencing of the respective areas of *KCNC2* (NM_139137) using the BigDye Terminator v3.1 Cycle Sequencing kit on an ABI3730XL DNA Analyzer to confirm the described mutations and define the inheritance model (Applied Biosystems; primer sequences available upon request).

### Protein structure analysis

No experimentally solved protein structure was available for the human K_V_3.2 channel. We generated a model of the protein structure using the RaptorX webserver^12^ that covered all 638 residues of the protein (NM_139137; NP_631875). Two scores were used for the identification of variant sensitive amino acid residues. First, we used our recently developed and validated score that identifies paralog conserved regions across genes of the same gene family (‘Para_zscore’)^13^. In a follow-up study, we showed that paralog conserved regions are enriched for patient variants^14^, in particular in neurodevelopmental diseases^13^. Secondly, we used the MTR-score that quantifies the constraint of each residue to missense variants in the general population. It was demonstrated that the most constraint regions are enriched for pathogenic variants in ClinVar and HGMD^15^. By combining the evidence of both scores, we detect critical regions (>5 consecutively amino acids) that are paralog conserved (Para_zscore > 0) and constraint for missense variation (MTR <0.459). Variants and critical regions were visualized in PyMOL^16^.

### Functional investigations

Four variants were selected for functional analysis according to the associated phenotype, the location and the predicted impact to the structure of the protein. *Backbone and RNA preparation*. Vector pcDNA3.1 (+) + insert *KCNC2* WT and the mutant clones (NM_139137: c.375C>G/p.Cys125Trp/C125W; c.404A>G/p.Glu135Gly/E135G; c.656T>C/p.Phe219Ser/F219S; c.1309A>G/p.Thr437Ala/T437A) were acquired from GenScript USA Inc. WT and mutant cDNA sequences were fully re-sequenced before being used in experiments to confirm the variant and exclude the presence of any additional sequence alterations. cRNA was prepared using the SP6 mMessage kit from Ambion according to the manufacturer’s instructions.

#### Electrophysiology

Collagenase-treated *Xenopus laevis* oocytes were acquired from Ecocyte Bioscience, Dortmund, Germany (1 mg/ml type CLS II collagenase, Biochrom, Berlin, Germany in OR-2 solutionin mM: 82.5 NaCl, 2.5 KCl, 1 MgCl_2_ and 5 HEPES, pH7.5), washed three times and stored at 16°C in Barth solution (in mM: 88 NaCl, 2.4 NaHCO_3_, 1 KCl, 0.33 Ca(NO_3_)_2_, 0.41 CaCl_2_, 0.82 MgSO_4_ and 5 Tris-HCl, pH7.4 with NaOH) supplemented with 50 µg/ml gentamicin (Biochrom). To compare current amplitudes of WT and mutant channels, 70 nl of cRNA encoding WT or mutant *KCNC2* cRNA (c = 1 µg/µl) were injected on the same day using the same batch of oocytes. For recordings of homozygous conditions of the WT or mutant K_V_3.2 channels 70 nl of cRNA (1.0) were injected. To be able to record the heterozygous condition, 35 nl of the WT and 35 nl of the related mutant *KCNC2* cRNA were injected. Injection was performed with the automated Roboinject system (Multi Channel Systems, Reutlingen, Germany), and oocytes were incubated for 5 days (at 17 °C) before the experiments were performed. Potassium currents in oocytes were recorded at room temperature (20–22 °C) using Roboocyte2 (Multi Channel Systems). For two-electrode voltage-clamp (TEVC) recordings, oocytes were impaled with two glass electrodes (resistance of 0.4–1MΩ) containing a solution of 1 M KCl and 1.5 M potassium acetate and clamped at a holding potential of −80 mV. Oocytes were perfused with an ND96 bath solution containing (in mM): 93.5 NaCl, 2 KCl, 1.8 CaCl_2_, 2 MgCl_2_ and 5 HEPES (pH7.6). Currents were sampled at 2 kHz.

#### Western blot analysis

For protein blotting, injected *Xenopus laevis* oocytes were lysed in a buffer containing 20mM Tris HCl (pH7.6), 100 M NaCl, 1% Triton X-100 and 1X complete protease inhibitors (Roche). After measuring the protein concentrations (BCA Systems, Thermo Fisher Scientific), 50 µg of protein were separated by SDS-PAGE on 8% polyacrylamide gels. Proteins were transferred onto polyvinylidene fluoride (PVDF) membranes (PALL Corporation) and protein blotting was performed using a monoclonal mouse antibody (S410-17) against K_V_3.2 (*KCNC2*) (ThermoFischer -MA5-27683) with a concentration of 1:500. Water-injected oocytes were used as negative control.

#### Data and statistical analysis

Data analysis and graphical illustrations were achieved using Roboocyte2+ (Multichannel Systems, Germany), Excel (Microsoft, USA) and GraphPad Prism Software (GraphPad Software, USA). Normality was tested using the Shapiro-Wilk test and statistical evaluation for multiple comparisons was conducted using one□way ANOVA on ranks with Dunn’s post hoc test. Statistical testing was performed with SigmaPlot 12.0 (Systat Software, Inc.) and di□erences between groups were considered significant with *p < 0.05. Data are reported as mean ± SEM (standard error of the mean).

## Results

### Genetic findings

We present 25 unique heterozygous missense variants identified in 27 patients including a first initial case published already by our group^17^. 22/25 variants were not found in control cohorts before. The remaining three variants were found once in the control cohort gnomAD (D128E, D194E, G200V). The presented *KCNC2* variants were grouped into three different categories (see also table 1). The first category (group 1: strong pathogenic variants, 10/27) are patients with *de novo* variants. Additionally, we acquired variants absent in large population databases (except D128E once described in gnomAD) that were either inherited from an unaffected parent (n=4) or variants where a positive family history could be determined without knowledge available about the inheritance model (n=4). We defined those as mild pathogenic or modifying variants (8/27, group 2) since minimum three out of four prediction scores indicate them as potentially damaging and the CADD score was high (>20). In 9/27 (group 3) cases the mode of inheritance was unclear. Since in those the prediction scores demonstrate either benign or tolerate status and the CADD score was low (<13) we declared these as variants of uncertain significance (see supplementary table).

**Table 1:** Detailed clinical and genetic information of the analyzed cohort. Variants in *italic*: functionally measured. *General information*: ASD autism spectrum disorder, dn *de novo*, f female, FH p/n family history positive or negative, ID intellectual disability, m male, mi maternal inherited, mc maternal non-affected carrier, N normal, NA not available, pc paternal non-affected carrier, Pt. patient. *Seizures and epilepsies*: Abs absences, CAE childhood absence epilepsy, CSWS continuous spike and wave during sleep, DEE developmental and epileptic encephalopathy, EOAE early onset absence epilepsy, FE focal epilepsy, FS febrile seizures, G generalized, GGE genetic generalized epilepsy, GTCS generalized tonic-clonic seizure, IS infantile spasms, JAE juvenile absence epilepsy, JME juvenile myoclonic epilepsy, LGS Lennox-Gastaut syndrome, Myo myoclonic seizures, SE status epilepticus, T-B temporal bilateral, T-L temporal left, TLE temporal lobe epilepsy, WS West syndrome. *Antiepileptic drugs (AED) and other treatments*: CZP clonazepam, ESM ethosuximide, KD ketogenic diet, LEV levetiracetam, OXC oxcarbazepine, P pharmaco-resistant, PB phenobarbital, sf seizure free, TPM topiramate, VPA valproic acid, ZNS zonisamide.

| Pt. No. | Protein variant | Inheritance mode (FH p/n) | f/m | syndrome | Seizure types | Neurological examination | ID/developmental delay | EEG | cMRI | Outcome (Sf Treatments) |
| --- | --- | --- | --- | --- | --- | --- | --- | --- | --- | --- |
| Group1: strong pathogenic variants (*de novo*), 10/27 |  |  |  |  |  |  |  |  |  |  |
| 1 | C125W | Dn | m | EOAE | Abs, Myo, GTCS | Facial dysmorphism, hypotonia, ataxia, ASD | mild-moderate | G | N | Sf (VPA, CZP) |
| 2 | E135G | Dn | m | DEE | Myo, atonic | N | mild | G | N | P |
| 3 | D167Y<sup>15</sup> | Dn | f | DEE: WS to LGS | IS, atypical Abs, Myo, focal unaware, GTCS | ataxic gait, hypotonia, speech disturbance, macrocephaly | mild-moderate | hypsarrhythmia, later multifocal | arachnoidal cysts left temporo-basal, progressive brain atrophy | P |
| 4 | F219S | Dn (p- Brother IGE with absences) | f | GGE | GTCS | N | no | G | N | P |
| 5 | R351K | Dn | m | DEE | GTCS, Myo, focal unaware | N | mild | CSWS | N | Sf (VPA) |
| 6 | R351K | Dn (p- brother with rolandic epilepsy and normal development) | m | DEE | GTCS, Myo, Abs, focal unaware | ASD, hyperactivity, impaired sleep, aphasia | severe | CSWS | N | P |
| 7 | F382C | Dn | m | GGE (JME) | Myo, atonic, clonic | N | no | G | venous anomaly | P |
| 8 | T437A | Dn | m | EOAE | Abs, tonic | Averbal and not follow commands | profound | G | N | P |
| 9 | T437N | Dn | f | DEE | FS, Myo | N | moderate | G | N | P |
| 10 | T437N | Dn | m | DEE | FS, Myo, Abs | hyperactivity | moderate | G | N | P |
| Group 2: mild pathogenic or modifying variants (8/27) |  |  |  |  |  |  |  |  |  |  |
| 11 | T32A | Mc(n) | f | GGE | GTCS, Abs | N | no | N | NA | Sf (VPA) |
| 12 | D128E | NA (n) | f | GGE | Abs, GTCS | N | no | G | generalized brain atrophy | Sf (VPA, ESM) |
| 13 | D144E | NA (p–maternal uncle) | m | GGE (JAE) | GTCS, Abs | N | no | G | NA | P (sf 2y VPA) |
| 14 | V330M | Mc (n) | f | GGE | GTCS | N | mild | G | N | Sf (VPA) |
| 15 | S333T | Mc (n) | f | Dravet-like (DEE) | GTCS, Myo, Abs | N | mild | G | N | Sf (VPA) |
| 16 | I465V | NA (p – father and first cousin had febrile convulsions) | m | FE | FS, SE, GTCS, focal aware and unaware | NA | NA | T-B | N | P |
| 17 | N530H | NA (n) | f | GGE | GTCS, Myo | depression and anxious | no | G | NA | P |
| 18 | S636F | Mc (p-GTCS in sister with proven gene mutation) | m | FE | Focal aware, GTCS | N | no | G | N | Sf (VPA) |
| Group 3: Variants of uncertain significance (9/27) |  |  |  |  |  |  |  |  |  |  |
| 19 | S93R | Pc (p–two brothers, son and daughters) | f | GGE | GTCS, Abs | N | no | G | Colloid cyst in the third ventricle | P |
| 20 | D178E | Mi (p-FS in mother) | f | FE | GTCS, FS, focal unaware | Deficit of long-term verbal memory and selective visual attention, depression | no | multifocal | N | P (sf2y LEV + VPA) |
| 21 | D194E | NA (n) | f | TLE (FE) | focal aware and unaware | depression | no | T-L | left mesial temporal sclerosis | Sf (surgery, LEV, CZP, ZNS) |
| 22 | G200V | Mc (p-focal epilepsy in childhood in older brother) | f | DEE | Focal aware, GTCS | Sleep disturbances, spastic quadriplegia, microcephaly, scoliosis | severe | multifocal | bilateral polymicrogyria | P |
| 23 | K204Q | Mc (p – maternal uncle with febrile seizures until 3y) | m | FS only | FS | N | no | multifocal | NA | Sf (none) |
| 24 | Splicing affected (c.G687+6T) | Mc (p-maternal uncle and his son(nephew) with FS) | f | DEE | Focal un-ware, GTCS | autism, self-harming behaviour, sleep disturbance | Moderate to severe | multifocal | cortical dysplasia T-L | Sf (KD, OXC, TPM) |
| 25 | I560M | NA (n) | f | FE | Focal aware, GTCS, atonic | visual field reduced to the left | mild | G, multifocal | double cortex (posterior), polymicrogyria, hippocampal sclerosis right | P |
| 26 | E608K (NM_139136) | NA (n) | m | EOAE | Abs | N | mild | G | N | P |
| 27 | R629H | Mc (n) | f | DEE | IS, GTCS | aphasia, ataxia, depression | no | Multifocal | N | P (sf PB, stopped because of side effects) |

Interestingly, two mutations have recurrently been identified in our cohort (R351K–two patients and T437N/A–three patients). All patients with recurrent mutations had a severe DEE and a very homogenous clinical phenotype concerning seizure onset, seizure types and developmental problems.

### Patients

We identified 27 patients carrying 25 different *KCNC2* variants (Table 1), 12 males and 15 females, and separated the patients into three categories regarding the potential pathogenicity of their respective *KCNC2* variants. Within these groups we could describe the following phenotypes:

### Pathogenic variants (*de novo*) -10/27 patients

#### Genetic generalized epilepsy (GGE)

2/10 patients presented with subtypes of GGE featuring myoclonic and generalized tonic-clonic seizures. EEG recordings showed typical generalized epileptic discharges. The neurological examination was normal in both patients. In the available cMRI (cranial magnetic resonance imaging) one patient presented a developmental venous anomaly which is a unspecific finding for epilepsy. Both patients are pharmacoresistant.

#### Early-Onset Absence Epilepsy (EOAE)

2/10 Patients had EOAE with dominant absences. Both showed normal MRI and, in the EEG, classical generalized epileptiform discharges. They suffered from mild ID. and demonstrated additional abnormalities including facial dysmorphism, hypotonia, ataxia, autism spectrum disorder. One of both patients was non-verbal. The other patient received seizure freedom using a combination of valproic acid (VPA) and clobazam.

#### Developmental and epileptic encephalopathy (DEE)

Furthermore, six patients with a DEE could be identified carrying a *de novo KCNC2* variant (6/10). ID was present in all patients with a moderate to severe intensity. Further neuro-psychiatric findings were ataxia, hypotonia, macrocephaly, autism, hyperactivity, sleep disturbance or problems with speech in 3/6 patients while the other three patients showed no neurological abnormality. Depending on the syndrome, the EEGs showed continuous electrical status epilepticus during slow wave sleep (CSWS) in two patients, multifocal or bilateral/generalized discharges as well as hypsarrhythmia. The cMRI was normal in 5/6 patients, while one patient showed an arachnoidal cyst combined with brain atrophy. Only one DEE patient achieved seizure freedom using VPA.

### Modifying variants -8/27 patients

#### Genetic generalized epilepsy (GGE)

5/9 patients presented with subtypes of GGE featuring absences, myoclonic and generalized tonic-clonic seizures. EEG recordings showed typical generalized epileptic discharges or normal interictal results. The neurological examination was normal in all patients. One patient with GGE suffered from psychiatric symptoms with depression and anxiety, one showed a mild intellectual disability (ID). In the available cMRI one patient presented a generalized brain atrophy, which is a unspecific finding for epilepsy. 4/5 GGE patients were seizure-free (one with transient effect over 2 years) using VPA in monotherapy or in combination.

#### Focal epilepsies (FE)

2/9 cases were suffering from FE. The Neurological examination and the cMRI was normal in one patient and in the other not available. None of them suffered from ID. EEG demonstrated bilateral temporal and generalized discharges. One patient achieved seizure freedom using VPA.

#### Developmental and epileptic encephalopathy (DEE)

Furthermore, one patient (1/8) with a DEE could be identified. This patient showed a mild ID but no other neuro-psychiatric findings. The EEG showed generalized discharges and the cMRI was normal. The patient received seizure freedom using VPA.

### Variants of uncertain significants -9/27 patients

#### Genetic generalized epilepsy (GGE)

1/8 patients presented with GGE featuring absences and generalized tonic-clonic seizures. EEG recordings showed typical generalized epileptic discharges and the neurological examination was normal. In the available cMRI the patient presented only a colloid cyst in the third ventricle, which is a finding unspecific for epilepsy. *Early-Onset Absence Epilepsy (EOAE)*. 1/8 Patients had EOAE with absences. The patient showed a normal MRI and classical generalized epileptiform discharges in the EEG. The Neurological examination was normal. The patient presented with a mild ID.

#### Focal epilepsies (FE)

3/8 cases were suffering from FE with structural findings in two patients (left mesial temporal sclerosis, posterior double cortex combined with polymicrogyria). The cMRI in the third patient was normal. Neuro-psychological deficits were present in all patients including depression, reduced visual field and long-term verbal memory deficit. One of them suffered from mild ID. EEG demonstrated left temporal, generalized or multifocal discharges. 2/3 patients achieved seizure freedom, one of them with VPA (only transient) and one received seizure freedom after epilepsy surgery.

#### Developmental and epileptic encephalopathy (DEE)

Furthermore, three patients (3/9) showed a DEE. ID was present in two patients, with a severe intensity. Further neuro-psychiatric findings were microcephaly, autism, quadriplegia, sleep disturbance, ataxia, depression or problems with speech. The EEGs showed multifocal discharges. In the cMRIs pathological findings were described like a cortical dysplasia or bilateral polymicrogyria. One DEE patient achieved seizure freedom using a combination of ketogenic diet, oxcarbazepine and topiramate, one under a medication of phenobarbital and one patient stayed pharmaco-resistant.

#### Febrile seizures (FS)

One patient (1/8) had FS only in early childhood, but no other seizure types and did not require medication. The patient had multifocal discharges on EEG, neuroimaging was not performed.

### Protein structure analysis

Based on the knowledge about potassium channel structures in general, we could locate our *KCNC2* variants across the six transmembrane segments of the K_V_3.2 subunit as well as in the long cytoplasmic N-and C-terminal regions^18^ (Figure 1A). The fourth transmembrane domain of the K_V_3.2 subunit builds a voltage sensor and the extracellular loop between the fifth and the sixth transmembrane domain serves as a selectivity filter for potassium ions. The identified variants were located in the N-and C-terminal part as well as in the last four transmembrane domains but the localization of each variant did not correlate to a specific phenotype. Since there was no crystal structure available, we modelled the structure of the K_V_3.2 subunit and mapped the identified variants in our model (Figure 1B). We identified exceptional protein regions, which are characterized by paralog conservation (Paraz-score) and depletion of population variants (MTR-score). 9/27 patient variants [F219S, V330M, S333T, R351K (two patients), F382C, T437A/N (three patients)] are located within these specific regions. They were all located close or within the transmembrane region of the protein. Interestingly, one further variant (I465V), was located in the transmembrane region. All others were localized in the cytoplasmic regions. Three variants were cytosolic N-terminal *de novo* variants (C125W, E135G, D167Y) within or close to the structured N-terminal cytoplasmic region, the so called T1 domain, which is important for the tetramerization of the protein^19^. The C-terminal region that harboured 5 patient variants is predicted to be mainly unstructured.

**Figure 1:**
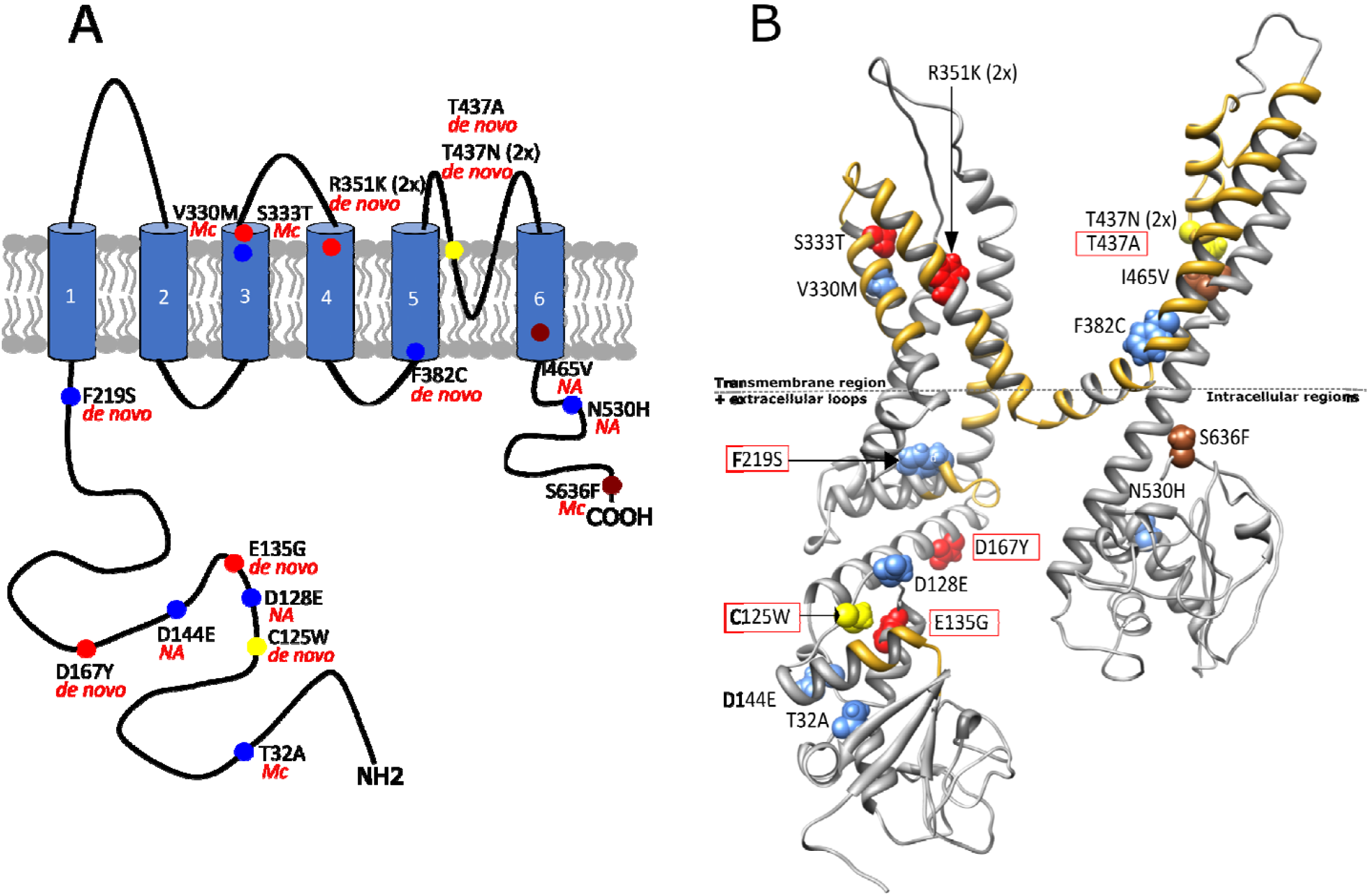
Structure of K_V_3.2 encoded by *KCNC2*. A. Schematic structure of K_V_3.2 subunit. The subunit consists of 6 transmembrane segments (1-6) with long C-and N-terminal regions. The N-terminal plays a crucial role for the tetramerization of the channel. The 4^th^ transmembrane segment works as the voltage sensor and the extracellular loop between the 5^th^ and the 6^th^ transmembrane segment forms the selectivity filter for K^+^ ions. Variants are color-coded according to the phenotype of the patient: red-developmental and epileptic encephalopathy (DEE), yellow-early onset absence epilepsy (EOAE), blue-genetic generalized epilepsy (GGE), brown-focal epilepsy (FE) and grey-febrile seizure (FS). B. The three-dimensional structure of K_V_3.2 predicted by RaptorX with *KCNC2* variants and phenotypes included (abbreviations see above). The golden areas within the structure are highly conserved regions characterized by paralog conservation (Paraz-score) and depletion of population variants (MTR-score). Extracellular loops are shown above the dotted line, below the line the intracellular N-and C-terminal regions are shown. The splice variant and E608K are not shown within the structure. E608K is only expressed in transcript number NM_139136 and not on NM_139137 which was used to create the structure. Red-rimmed variants were selected for functional analysis either measured here or previously described by ourselves.^*17*^

In summary, we could not detect a strong phenotype-localization correlation in the predicted structure of the K_V_3.2 subunit, but most of the identified variants are located in specific relevant and conserved areas of the channel.

### Functional analysis

We performed the two-electrode voltage clamp technique using the *Xenopus laevis* oocyte expression system and recorded the currents of four different variants which were selected according to the location of the variant within the channel structure as well as the phenotype of the patient (C125W-EOAE, E135G-DEE, F219S-GGE and T437A-EOAE, see also Figure 1). All of them showed a *de novo* inheritance pattern and none of them was found in control cohorts (gnomAD). C125W and E135G are located within the T1 domain which is important for the tetramerization of the channel (see Figure 1A). In this region, the Zn^2+^ coordinating motif is also located (Hx_5_Cx_20_CC) which is important in bridging the interaction interface between two proteins and stabilize the tetrameric protein structure. Additionally, the Zn^2+^location within the Shaw and Shaker family is different suggesting that zinc may play a role in differentiating Shaw from Shaker T1 in assembly^19^. F219S is located shortly before the first transmembrane domain and T437A is located within the P-domain (TxxT/SxGY/FG), which acts as the K^+^ selectivity filter, and affects the second important amino acid of the P-domain motif^20^.

The functionally analysed variant F219S-GGE showed a complete loss-of-function in the homo-and heterozygous state with current amplitudes comparable to the ones obtained from water injected control oocytes (Figure 3A). Thus, this variant causes a dominant negative effect on WT channels (Figure 3A/3C). T437A-EOAE lead to a significant reduction of the current amplitude, while C125W-EOAE showed a significant increase in the current amplitude. The analysis of E135G-DEE could not demonstrate any differences compared to oocytes injected with the WT subunit (Figure 2A and 2B). To make sure that all variants have been expressed in the injected oocytes we performed a western blot analysis. This analysis showed a band at about 90 kDa in all protein lysates except for the water injected control oocytes. As loading control, the housekeeping gene Beta-actin was used, and all protein lysates showed a band at about 40 kDa. Thus, all variants showed an expression in oocytes (Figure 2D and 3B). The analysis of the channel kinetic of the variants C125W-EOAE, E135G-DEE, T437A-EOAE, showed a significant shift of the activation curve to more hyperpolarized potentials (Figure 2E) and in total a slower deactivation compared to oocytes expressing WT channels (Figure 2F). E135G-DEE showed a significantly slower deactivation time constant at all recorded voltages except for 0 mV compared to cells injected with RNA encoding only WT subunits. For C125W-EOAE and T437A-EOAE the deactivation time constant was significantly reduced between −50 mV and −20 mV or −30 mV respectively in comparison to oocytes expressing WT channels (Figure 2F). Thus, the analysis of the variants C125W, E135G and T437A in total demonstrates a gain-of-function in the channel kinetics. Additionally, the membrane potential of the variants C125W, F219S and T437A was significantly shifted to more hyperpolarized potentials (Figure 2C and 3D).

**Figure 2:**
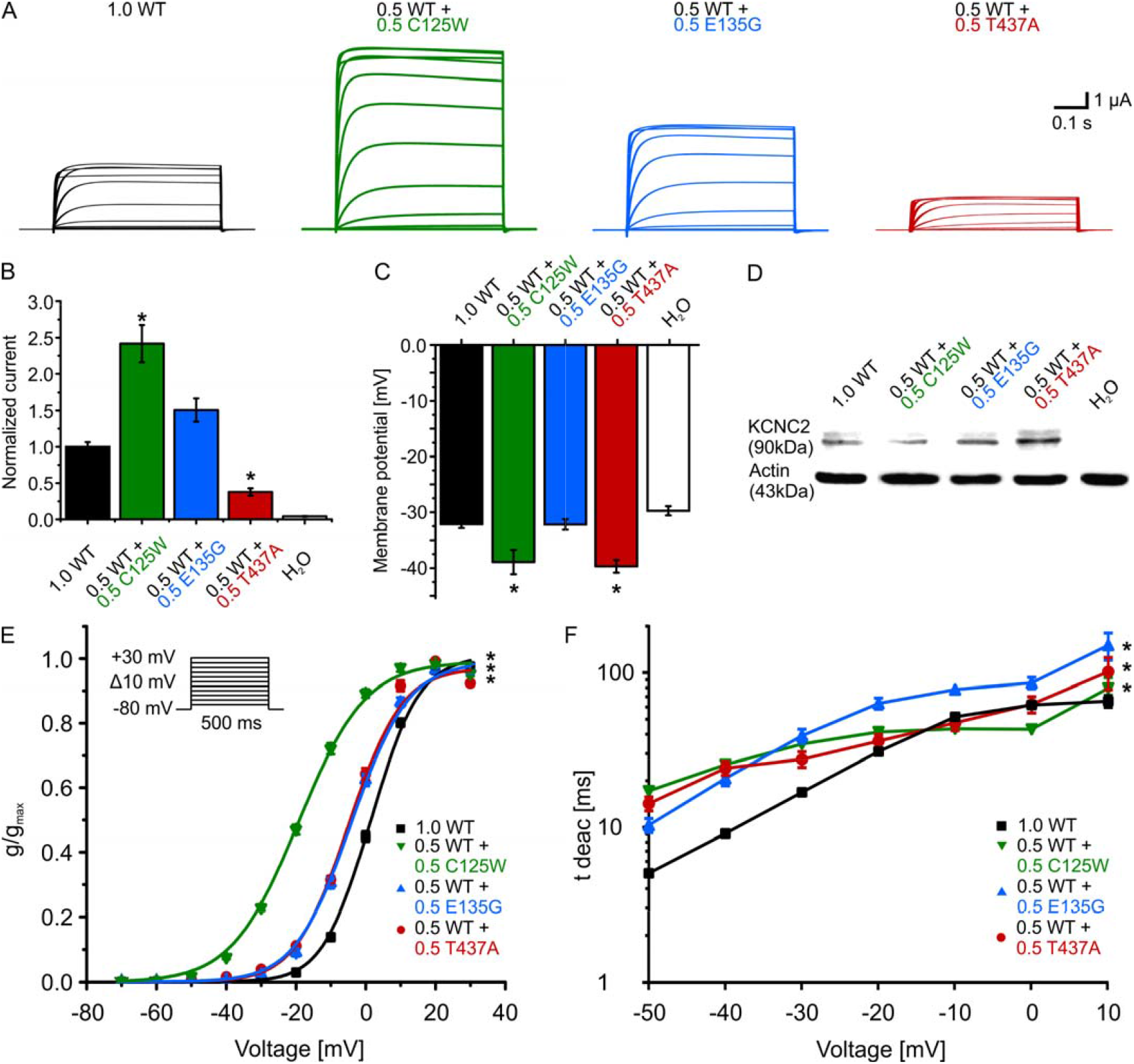
Electrophysiological analysis of selected *KCNC2* variants. Functional analysis of the variants C125W, E135G and T437A compared to wild type (WT). The more severe phenotypes C125W-EOAE (increased current amplitude/activation at more hyperpolarized potentials/slow deactivation), E135G-DEE (normal current/activation at more hyperpolarized potentials/slow deactivation) and T437A-EOAE (reduced current amplitude/activation at more hyperpolarized potentials/slow deactivation) demonstrate gain-of-function. A. Representative traces of K_V_3.2 currents recorded in *Xenopus laevis* oocytes expressing wild type (WT) or the different variants (C125W, E135G, T437A) in response to voltage steps from −70 mV to +30 mV (with an increment of 10 mV). B: Mean current amplitudes of oocytes injected with WT (n=101), and equal amounts of WT + C125W (n=40), WT + E135G (n=31), WT + T437A (n=41) or water (1.0, n=44). C. Resting membrane potentials of oocytes injected with WT (n=101) and equal amounts of WT + C125W (n=40), WT + E135G (n=31), WT + F219S (n=29), WT + T437A (n=41) or water (n=44). Shown are means ± SEM (standard error of the mean). Statistically significant differences between WT channels and the tested groups were verified by ANOVA on ranks (indicated by asterisks). D.Immunoblot analysis for lysates of *Xenopus laevis* oocytes injected with cRNA for K_V_3.2 WT and equal amounts of WT + C125W, WT + E135G, WT + T437A or water. All channels showed a band at about 90 kDa. E.Mean voltage-dependent activation of K_V_3.2 channel for WT (n = 101), equal amounts of WT+C125W (n=40), WT+E135G (n=31) and WT+T437A (n=42). Lines illustrate Boltzmann Function fit to the data points. All activation curves showed a significant shift to more hyperpolarized potentials in comparison to WT channel alone. All data are shown as means ± SEM. F.Mean voltage-dependent deactivation time constant of K_V_3.2 channel WT (n=72), WT+C125W (n=40), WT+E135G (n=12) and WT+T437A (n=20). All deactivation curves showed a significantly slower deactivation in comparison to channels only containing WT subunit. All data are shown as means ± SEM.

**Figure 3:**
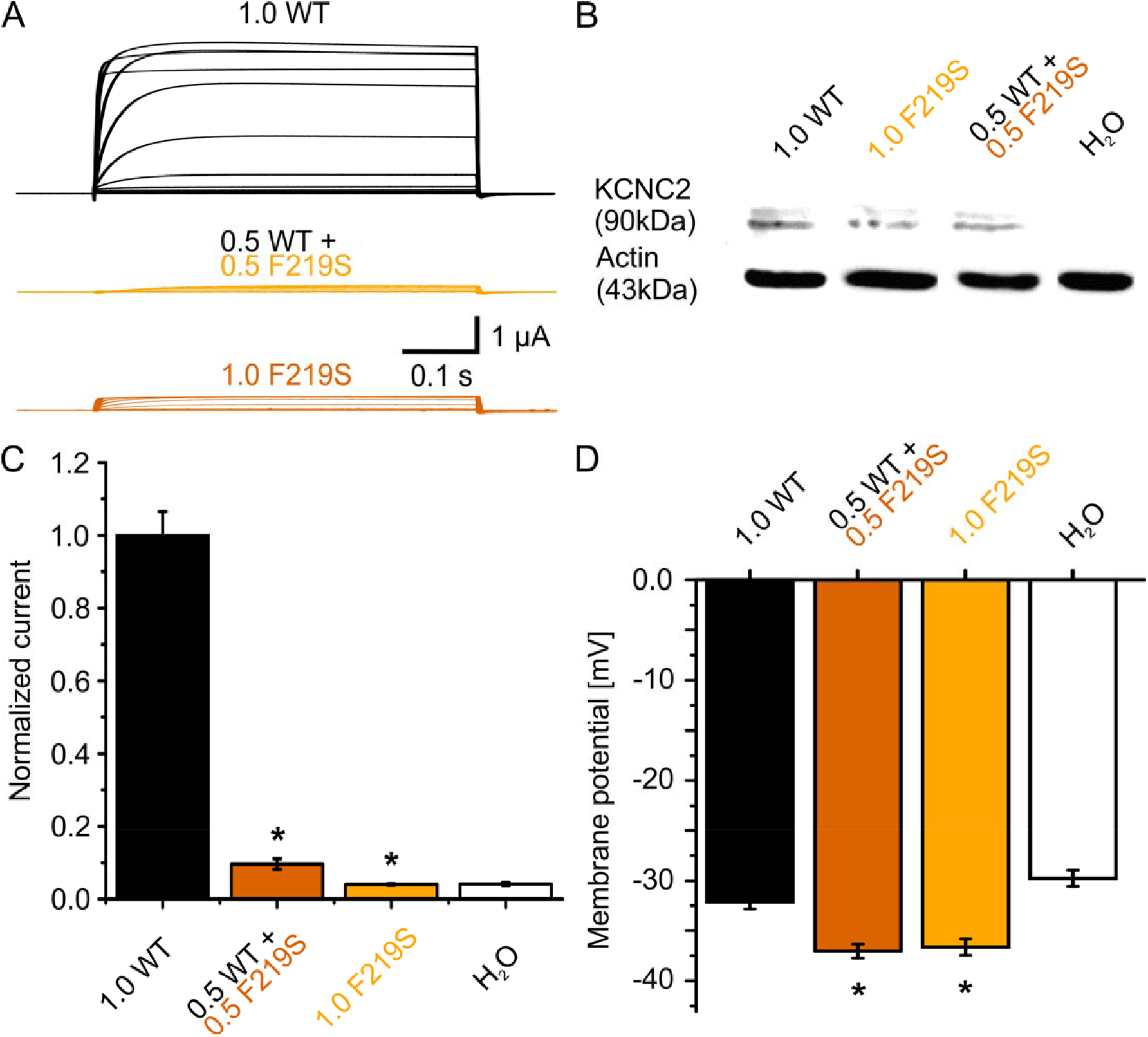
Electrophysiological analysis of the p.F219S *KCNC2* variant. Functional analysis for the variant F219S compared to wild type (WT). In summary the figures show that the milder phenotype F219S-GGE has a dramatically dominant negative effect in a sense of a loss-of-function. A. Representative traces of K_V_3.2 currents in *Xenopus laevis* oocytes expressing WT, F219S or a 1:1 mixture of both in response to the voltage steps from −70mV to + 30mV. B. Immunoblot analysis for lysates of *Xenopus laevis* oocytes injected with cRNA for K_V_3.2 WT, F219S, equal amounts of WT+F219S or water. All channels showed a band at about 90kDa. C. Mean current amplitudes of analyzed oocytes injected with WT (n=101), F219S (n=39), equal amounts of WT + F219S (n=29) or water (n=44). D.Resting membrane potentials of oocytes injected with WT (n=101), F219S (n=39), equal amounts of WT + F219S (n=29) or water (n=44). Shown are means ± SEM (standard error of the mean). Statistically significant differences between WT channels and the tested groups were verified by ANOVA on ranks (indicated by asterisks).

Taken together the variant T437A-EOAE showed a mixed effect on the channel function, a gain-of-function in the channel kinetic and a loss-of-function in the normalized current amplitude. For the other two variants (C125W-EOAE and E135G-DEE), the gain-of-function was the predominant effect.

In summary, the more severe phenotypes demonstrate a clear gain-of-function effect, whereas the milder phenotype F219S-GGE causes a dramatic dominant negative loss-of-function effect.

## Discussion

We describe *KCNC2* as a novel genetic etiology for human epilepsies. Phenotypes range from relatively mild generalized epilepsies to severe developmental and epileptic encephalopathies related to specific functional changes.

The variants collected demonstrate a spectrum of severity. Therefore, we divided these into three different categories. The first category includes only *de novo* mutations that are disease-causing (10/27 variants). No other relevant mutations in other known epilepsy genes were found in these patients. The second category includes patients with variants in *KCNC2* which are likely to be disease causing or have only a modifying effect since the prediction scores indicated pathogenicity, but the variants were inherited by unaffected family members or the inheritance mode was unclear (8/27). The third category contains variants of uncertain significance (9/27). The clinical spectrum observed in *KCNC2*-related disorders is very broad concerning the extent of severity. DEE was the main phenotype (37% in total cohort and in group 1&2 variants: 39%), but closely followed by GGE (30% in total cohort and in group1&2 variants: 39%), FE (19% in total cohort and in group 1&2 cohort 11%), EOAE (11% in total cohort and in group 1&2 variants 11%) and pure febrile seizures in one case (variant of uncertain significance). Nevertheless, the recurrent variants (R351K and T437N) had a recognizable homogenous clinical picture hinting a unique genotype-phenotype correlation.

The GGE-*KCNC2* cases (group 1&2 variants) demonstrated a reduced response to antiepileptic medication since only 57% were pharmaco-responsive compared to approximately 90% in the general GGE population^21^. Interestingly, 8/18 patients became seizure-free (variants group 1&2) and all of them using valproic acid (VPA) as monotherapy or in combination. The patients responding to VPA carry variants which mainly cluster in two regions. One is the intracellular N-terminal part including the variants (T32A, D128E, D144E) and the other one the extracellular region of the third and fourth domain including the variants V330M, S333T and R351K. VPA is an antiepileptic drug with a broad spectrum of mechanisms of actions. VPA has been demonstrated to limit high-frequency repetitive firing in cultured neurons^22^. This effect is linked to the modulation of sodium, calcium and potassium channels, especially with a use-dependent decrease in inward sodium currents. Additionally, VPA increases the amplitude of the late potassium outward currents, further increasing the threshold for epileptiform activity^22^ which might explain the special effect of this drug in our cohort.

Our cohort includes also patients with structural brain changes: relevant structural changes were found in 15% of our cases (4/27, mesial temporal sclerosis, polymicrogyria, cortical dysplasia, double cortex associated with polymicrogyria). All of them carried a variant of uncertain significance. Therefore, it is unclear whether variants in the *KCNC2* gene may affect brain development and can also induce structural changes, as known, for example, for mutations in the *GPR56, DCX* or *LIS1* genes^20,23,24^ or is only a modifying factor in these cases. Nevertheless, associated brain malformations were described before for other ion channel genes like *SCN1A* and *SCN3A*^25,26^.

*KCNC2* has previously been proposed as a modifying factor^17^, and to play a role in other neuro-psychiatric diseases such as ataxia^27,28^, schizophrenia^29^, bipolar disorder^30^, and other DEEs^31^. Our patients did also display additional neurological features including facial dysmorphism, ataxia, speech disturbance, depression, hyperactivity and autism spectrum disorder with additional support to this hypothesis. Additionally, we could demonstrate a much broader clinical spectrum including GGE, EOAE and FE. Our collected variants can be dedicated as either strong disease causing, mild pathogenic or characterized as phenotype modifiers.

The virtual structure of K_V_3.2 identified very important regions, which are characterized by paralog conservation (Paraz-score) and depletion of population variants (MTR-score). We could not find a strong phenotype-structure association, but the variants of our patients were concentrated at the C-terminus, N-terminus and transmembrane segments 3 to 6 indicating a high relevance of these regions to channel function.

We selected four variants for functional analysis and selected them based on the phenotype of the patients as well as the location within the channel. The functional results demonstrated a gain-of-function in the more severe phenotypes DEE and EOAE, whereas in the GGE associated variant a dramatic loss-of-function effect was observed. Among all K_V_ channels, channels of the K_V_3 family in particular play a crucial role in the rapid repolarization of action potentials and therefore dictate action potential duration. The K_V_3.2 subunit is predominantly expressed in the brain specifically in parvalbumin-as well as somatostatin-expressing GABAergic interneurons in deep cortical layers^5,32^ and therefore responsible for the modulation of excitation. Furthermore, the functional analysis demonstrated that voltage-gated potassium channels are important for the resting membrane potential and the regulation of firing, action potential duration and neurotransmitter release^5,6^. The K_V_3.2 subunit has some unusual electrophysiological properties like the fast rate of deactivation upon repolarization. This rate is significantly faster than that of any other known neuronal voltage-gated K^+^ channel. Thus, K_V_3.2 plays a crucial role in fast-spiking interneurons^33^. It is therefore reasonable that inhibitory interneurons play an important role in the development of epileptic seizures in the cases described here. In many generalized epilepsies, inhibitory interneurons play an essential role, for example in Dravet syndrome and GEFS+ which are associated to pathogenic variants in *SCN1A* encoding the main Na^+^ channel in inhibitory neurons^34–36^ or *KCNC1* mutations in progressive myoclonic epilepsy^8^.

K_V_3.2^-/-^ knockout mice presented with specific changes in their cortical EEG patterns and additionally showed increased susceptibility to epileptic seizures^37^. This again emphasizes how important it is for inhibitory interneurons to be able to generate high-frequency firing so that balanced cortical operations can occur. As described for other variants in potassium channel genes associated with epilepsy^38^, the recorded K_V_3.2 variants demonstrate gain-and loss-of-function effects.

In conclusion, *KCNC2* is a novel and important gene for a broad spectrum of epilepsy syndromes with an interesting phenotype-pathophysiology correlation and a potential concept for precision medicine.

## Supporting information

KCNC2 Manuscript

## Data Availability

I hereby confirm that all data referred to the manuscript are available.

## Supplemental information description

**Supplmentary table**: Analysis of the detected *KCNC2* variants (**NM_139137)** by different prediction tools. Protein variants in bold were functionally analyzed. Cadd phred of 20 or more marked in bold.

## Acknowledgements

We thank the participants and their family members for taking part in the study. The study received support through the German Research Foundation (HE5415/5-1, HE5415/6-1 to I.H., and WE4896/3-1 to Y.W., KU 911/21-2 and KU 911/22-1 to W.S.K.) and the DFG/FNR

INTER Research Unit FOR2715 (We4896/4-1 to Y.W, He5415/7-1 to I.H., INTER/DFG/17/11583046 to P.M.). N.S., T.B., D.L., P.M., and Y.W. were supported by the BMBF Treat-ION grant (01GM1907). I.H. was supported by The Hartwell Foundation through an Individual Biomedical Research Award. This work was also supported by the National Institute for Neurological Disorders and Stroke (K02 NS112600), including support through the Center Without Walls on Ion channel function in epilepsy (“Channelopathy-associated Research Center”, U54 NS108874), the Eunice Kennedy Shriver National Institute of Child Health and Human Development through the Intellectual and Developmental Disabilities Research Center (IDDRC) at Children’s Hospital of Philadelphia and the University of Pennsylvania (U54 HD086984), and by intramural funds of the Children’s Hospital of Philadelphia through the Epilepsy NeuroGenetics Initiative (ENGIN). Research reported in this publication was also supported by the National Center for Advancing Translational Sciences of the National Institutes of Health under Award Number UL1TR001878. The content is solely the responsibility of the authors and does not necessarily represent the official views of the NIH. This project was also supported in part by the Institute for Translational Medicine and Therapeutics’ (ITMAT) Transdisciplinary Program in Translational Medicine and Therapeutics at the Perelman School of Medicine of the University of Pennsylvania.

The study also received support through the EuroEPINOMICS-Rare Epilepsy Syndrome (RES) Consortium as well as) and by the Genomics Research and Innovation Network (GRIN, grinnetwork.org). I.H. also received support through the International League Against Epilepsy (ILAE). H.M. was supported by intramural funds of the Christian-Albrechts-University of Kiel. PS and FZ developed this work within the framework of the DINOGMI Department of Excellence of MIUR 2018-2022 (legge 232 del 2016).

The DECODE-EE project (Health Research Call 2018, Tuscany Region) provided research funding to RG and AV.

HK and MD were supported by a research grant from Science Foundation Ireland (SFI) under Grant Number 16/RC/3948 and co-funded under the European Regional Development Fund and by FutureNeuro industry partners.

The ITAUBG centre thanks Dr Pippucci for the supervision in genetic data analysis, the Neurogenetics Laboratory staff (led by Prof. Carelli V) and all the physicians and nurses of the Epilepsy Centre from the Bellaria Hospital in Bologna. The effort was supported by the “Ricerca Corrente” funding from the Italian Ministry of Health.

We thank the Epi25 principal investigators, local staff from individual cohorts, and all of the patients with epilepsy who participated in the study for making possible this global collaboration and resource to advance epilepsy genetics research. This work is part of the Centers for Common Disease Genomics (CCDG) program, funded by the National Human Genome Research Institute (NHGRI) and the National Heart, Lung, and Blood Institute (NHLBI). CCDG-funded Epi25 research activities at the Broad Institute, including genomic data generation in the Broad Genomics Platform, are supported by NHGRI grant UM1 HG008895 (PIs: Eric Lander, Stacey Gabriel, Mark Daly, Sekar Kathiresan). The Genome Sequencing Program efforts were also supported by NHGRI grant 5U01HG009088-02. The content is solely the responsibility of the authors and does not necessarily represent the official views of the National Institutes of Health. We thank the Stanley Center for Psychiatric Research at the Broad Institute for supporting the genomic data generation efforts and control sample aggregation.

## Author contributions

NS worked on the functional analysis of the variants and wrote the manuscript together with SS, MP, AR, IH and YW.

SS performed genetic and functional analysis and wrote the manuscript together with NS, MP, AR, IH and YW.

UBSH re-analyzed the functional data and edited the manuscript.

MP worked on the clinical data of the patients, performed genetic analysis and wrote the manuscript together with NS, SS, AR, IH and YW.

AR performed the detail clinical analysis of the patients and wrote the manuscript together with NS, SS, MP, IH and YW.

TB performed the crystal structure of K_V_3.2. together with DL and PM.

PBA, HB, AB, FB, RJB, BZB, MGD, RG, GH, MI, HK, KMK, IK, WSK, HL, LL, EL, RM, MM, SM, LM, RO, BO, SSP, FR, TG, PSR, FR, AR, PS1, PS2, GAT, AV, FZ1, FZ2 performed detailed clinical evaluation and further genetic analysis.

DL and PM performed additional genetic analysis, performed the virtual crystal structure of K_V_3.2 and edited the manuscript.

HM planned the study together with IH and YW, performed clinical analysis of the patients and edited the manuscript.

IH planned the study together with YW, supervised the analysis and edited the manuscript. YW planned and coordinated the study, supervised the analysis, wrote and edited the manuscript together with IH, NS, SS, MP and AR.

