## Supplementary material for "Heterozygous variants in *KCNC2* cause a broad spectrum of epilepsy phenotypes associated with characteristic functional alterations": KCNC2 Manuscript

**Supplmentary table**: Analysis of the detected *KCNC2* variants (**NM_139137)** by different prediction tools (marked in bold if deleterious, damaging, possibly damaging or disease causing). Protein variants in bold were functionally analyzed. Cadd phred of 20 or more marked in bold.

| **Pt. No** | **Mutations** | **Protein variants** | **SIFT (T, tolerated; D, deleterious)** | **Polyphen2 HVAR (B, benign, P; possibly damaging; D, probably damaging)** | **Polyphen HDIV (B, benign, P; possibly damaging; D, probably damaging)** | **MutationTaster (N, neutral; D, disease causing)** | **CADD phred** |
| --- | --- | --- | --- | --- | --- | --- | --- |
| **Group 1: strong pathogenic variants (*de novo*), 10/27** | | | | | | | |
| 1 | c.C375G | **p.C125W** | **D** | **D** | **D** | **D** | **26.9** |
| 2 | c.A404G | **p.E135G** | T | **D** | **D** | **D** | **27.4** |
| 3 | c.G499T | p.D167Y | T | **D** | **P** | **D** | **29.5** |
| 4 | c.T656C | **p.F219S** | **D** | **P** | **D** | **D** | **28.2** |
| 5/6 | c.G1052A | p.R351K | **D** | **D** | **D** | **D** | **27.8** |
| 7 | c.T1145G | p.F382C | **D** | **D** | **D** | **D** | **25.1** |
| 8 | c.A1309G | **p.T437A** | **D** | **D** | **D** | **D** | **24.7** |
| 9/10 | c.C1310A | p.T437N | **D** | **D** | **D** | **D** | **25.6** |
| **Group 2: mild pathogenic or modifying variants, 8/27** | | | | | | | |
| 11 | c.A94G | p.T32A | **D** | **P** | **D** | **D** | **25.1** |
| 12 | c.C384A | p.D128E | T | **P** | **D** | **D** | **27.8** |
| 13 | c.C432G | p.D144E | **D** | **P** | **P** | **D** | **24.2** |
| 14 | c.G988A | p.V330M | T | **P** | **P** | **D** | **24.3** |
| 15 | c.G998C | p.S333T | T | **P** | **D** | **D** | **24.1** |
| 16 | c.A1393G | p.I465V | T | **D** | **D** | **D** | **22.9** |
| 17 | c.A1588C | p.N530H | **D** | **P** | **P** | **D** | **21.4** |
| 18 | c.C1907T | p.S636F | **D** | **P** | **D** | **D** | **28.2** |
| **Group 3: variants of uncertain significance, 9/27** | | | | | | | |
| 19 | c.C279G | p.S93R | T | B | B | N | 2.744 |
| 20 | c.C534A | p.D178E | T | B | B | **D** | 10.89 |
| 21 | c.C582G | p.D194E | T | B | B | **D** | 10.12 |
| 22 | c.G599T | p.G200V | T | B | B | **D** | 7.611 |
| 23 | c.A610C | p.K204Q | T | B | B | **D** | 10.67 |
| 24 | c.G1822A (only in this transtript: NM_139136) | p.E608K | **D** | B | **P** | **D** | 12.91 |
| 25 | c.C1680G | p.I560M | T | B | B | **D** | 12.85 |
| 26 | c.G687+6T | intronic | - | - | - | - | - |
| 27 | c.G1886A | p.R629H | T | B | B | **D** | **23.6** |
